# Correlates of symptomatic remission among individuals with post-COVID-19 condition

**DOI:** 10.1101/2023.01.31.23285246

**Authors:** Roy H. Perlis, Mauricio Santillana, Katherine Ognyanova, David Lazer

**Affiliations:** Massachusetts General Hospital, Boston, MA; Harvard Medical School, Boston, MA; Northeastern University, Boston, MA; Rutgers University, New Brunswick, NJ

**Keywords:** SARS-CoV2, Long COVID, Post-acute sequelae of COVID-19, chronic, survey, cross-sectional study

## Abstract

**Importance:** Post-COVID-19 condition (PCC), or long COVID, has become prevalent. The course of this syndrome, and likelihood of remission, has not been characterized.

**Objective:** To quantify the rates of remission of PCC, and the sociodemographic features associated with remission.

**Design:** 16 waves of a 50-state U.S. non-probability internet survey conducted between August 2020 and November 2022

**Setting:** Population-based

**Participants:** Survey respondents age 18 and older

**Main Outcome and Measure:** PCC remission, defined as reporting full recovery from COVID-19 symptoms among individuals who on a prior survey wave reported experiencing continued COVID-19 symptoms beyond 2 months after the initial month of symptoms.

**Results:** Among 423 survey respondents reporting continued symptoms more than 2 months after acute test-confirmed COVID-19 illness, who then completed at least 1 subsequent survey, mean age was 53.7 (SD 13.6) years; 293 (69%) identified as women, and 130 (31%) as men; 9 (2%) identified as Asian, 29 (7%) as Black, 13 (3%) as Hispanic, 15 (4%) as another category including Native American or Pacific Islander, and the remaining 357 (84%) as White. Overall, 131/423 (31%) of those who completed a subsequent survey reported no longer being symptomatic. In Cox regression models, male gender, younger age, lesser impact of PCC symptoms at initial visit, and infection when the Omicron strain predominated were all statistically significantly associated with greater likelihood of remission; presence of ‘brain fog’ or shortness of breath were associated with lesser likelihood of remission.

**Conclusions and Relevance:** A minority of individuals reported remission of PCC symptoms, highlighting the importance of efforts to identify treatments for this syndrome or means of preventing it.

**Trial registration:** N/A

**Key Points:** *Question:* How often do individuals with post-COVID-19 condition, or long COVID, recover fully, and what predicts this recovery?

*Findings:* Among 423 individuals who initially reported post-COVID-19 condition and completed at least one subsequent survey, 131 (31%) later described symptomatic remission. Younger age, male gender, lesser symptom severity at initial survey, and infection during Omicron-predominance were associated with greater likelihood of reporting recovery.

*Meaning:* A minority of people with post-COVID-19 condition report spontaneous remission of symptoms, highlighting the importance of developing treatments for this syndrome.

## INTRODUCTION

Post-COVID-19 condition (PCC) describes the persistence of symptoms following acute Coronavirus-19 (COVID-19) illness^1^, at least 3 months after infection^2^. Studies using surveys^3,4,5^ or health records^6,7^ generally report point prevalences between 5 and 15%, depending on the definition and risk group. An emerging literature documents the substantial costs associated with PCC, or long COVID, in terms of functioning^8^, quality of life^9^, and economic impact^10^.

Multiple interventions are being investigated to address PCC, either in aggregate or in terms of specific symptoms^11^. However, little is known about the extent to which individuals will experience remission over time with or without treatment. A recent self-report study relying on daily symptom diaries found that about 15% of patients with PCC experienced remission after 12 months^12^. A smaller 12-month follow-up survey study estimated prevalence of remission of 9%^13^. Beyond acute symptoms, neither study investigated whether particular groups of individuals were less likely to experience remission. Understanding these characteristics of PCC could be critical in targeting interventions as they are developed, or identifying groups in which treatment may be particularly important.

To characterize the sociodemographic and clinical features associated with symptomatic remission of PCC, we used data from multiple waves of a 50-state US survey conducted approximately every 4-8 weeks since spring 2020, which includes questions about COVID-19 symptoms but does not focus solely on COVID-19, decreasing risk of selection bias toward individuals with a particular interest in the pandemic. Among those respondents who had previously reported PCC symptoms, we aimed to understand how often these symptoms resolved fully.

## METHOD

### Study Design

The generated a cohort using data from 16 waves of the COVID States Project, an academic consortium which has conducted a 50-state internet survey approximately every 4-8 weeks since spring 2020, with questions about illness duration beginning in August 2020 through November 2022. Eligible participants are age 18 and older, reside in the United States, and are able to provide written informed consent at the beginning of the survey. This non-probability sampled^14^ survey uses representative quotas to balance age, gender, race/ethnicity, and geographic distribution, ensuring representation of a sufficient number of participants in every state. The study has been classified as exempt by the Institutional Review Board of Harvard University. Results are described in accordance with AAPOR guidelines^15^.

### Measures

Prior COVID-19 infection was identified by asking respondents if and when they had received any positive COVID-19 test result, without specific language regarding type of test. Those who indicated a positive test were asked if symptoms had resolved; if they had not, respondents completed a checklist of common COVID-19 symptoms. They were also asked, on a 5-point scale, to identify how much their symptoms interfered with a normal day over the prior 2 weeks (not at all, a little, moderately, quite a bit, or extremely); a priori, this ordinal scale was dichotomized to reflect ‘less than moderately’ or ‘at least moderately’ for analysis. All respondents also provided sociodemographic information including age, gender, race and ethnicity, and zip code. Race and ethnicity were coded according to 5 United States Census categories and used to ensure representativeness of the US population in survey weighting.

To determine whether individuals were vaccinated prior to initial infection, month of vaccination was compared to first reported month of illness; we defined completion of primary series as 2 vaccinations before the first month of illness, or a single vaccination with the Johnson and Johnson vaccine. Predominant US COVID-19 variant at time of infection was estimated using CoVariants analysis of GISAID data^16^, according to the variant accounting for 50% or more of the total in a given month.

### Analysis

As in our prior work^5^, PCC was defined according to the World Health Organization^2^ description, encompassing individuals who endorsed continued symptoms on a survey whose start date was more than 2 months after the month in which they reported a positive COVID-19 test.

We defined the index visit as the first survey at which individuals reported long COVID. We then analyzed the last available survey for that respondent, and used survival analysis to account for variable follow-up interval.

We applied Cox regression in R 4.0^17^ to examine association of individual sociodemographic and clinical features with remission, after visual inspection and testing of Schoenfeld residuals to ensure that proportional hazards assumption was not rejected. To visualize results, we also generated Kaplan-Meier survival curves for a subset of variables significant in adjusted models.

## RESULTS

The cohort included 423 survey respondents who initially reported continued symptoms more than 2 months after acute test-confirmed COVID-19 illness, and then completed at least 1 subsequent survey, with mean age of 53.7 (SD 13.6); 293 (69%) identified as women, and 130 (31%) as men; 9 (2%) identified as Asian, 29 (7%) as Black, 13 (3%) as Hispanic, 15 (4%) as another category including Native American or Pacific Islander, and the remaining 357 (84%) as White. Additional characteristics of the cohort are summarized in Table 1. Overall, 131/423 (31%) of those who completed a subsequent survey reported that they were no longer symptomatic. Mean interval from baseline to last survey was 173 days (SD 150 days). (For comparison of those with follow-up visit to individuals who did not complete a subsequent survey wave, see Supplemental Table 1.)

**Table 1.** Characteristics of individuals who completed a subsequent survey after identifying as having long COVID symptoms on a prior one, contrasting those who did or did not report continued long COVID symptoms.

|  | Persistent | Remitted | Total (N=423) | P value |
| --- | --- | --- | --- | --- |
| **Gender (n %)** |  |  |  | .004 |
| Female | 215 (73.6%) | 78 (59.5%) | 293 (69.3%) |  |
| Male | 77 (26.4%) | 53 (40.5%) | 130 (30.7%) |  |
| **Respondent age (mean SD)** | 54.4 (13.2) | 52.3 (14.4) | 53.7 (13.6) | .14 |
| **Race and ethnicity** |  |  |  | .55 |
| Black | 16 (5.5%) | 13 (9.9%) | 29 (6.9%) |  |
| Asian American | 6 (2.1%) | 3 (2.3%) | 9 (2.1%) |  |
| Hispanic | 9 (3.1%) | 4 (3.1%) | 13 (3.1%) |  |
| Native American | 2 (0.7%) | 0 (0.0%) | 2 (0.5%) |  |
| Other | 7 (2.4%) | 3 (2.3%) | 10 (2.4%) |  |
| Pacific Islander | 3 (1.0%) | 0 (0.0%) | 3 (0.7%) |  |
| White | 249 (85.3%) | 108 (82.4%) | 357 (84.4%) |  |
| **US region** |  |  |  | .46 |
| Midwest | 86 (29.5%) | 37 (28.2%) | 123 (29.1%) |  |
| Northeast | 40 (13.7%) | 22 (16.8%) | 62 (14.7%) |  |
| South | 121 (41.4%) | 46 (35.1%) | 167 (39.5%) |  |
| West | 45 (15.4%) | 26 (19.8%) | 71 (16.8%) |  |
| **Impact of long COVID symptoms<sup>a</sup>** |  |  |  | .001 |
| Less than moderate | 116 (40.6%) | 75 (57.7%) | 191 (45.9%) |  |
| Moderate or greater | 170 (59.4%) | 55 (42.3%) | 225 (54.1%) |  |
| **Predominant COVID variant** |  |  |  | .27 |
| alpha | 21 (7.2%) | 3 (2.3%) | 24 (5.7%) |  |
| ancestral | 178 (61.0%) | 84 (64.1%) | 262 (61.9%) |  |
| delta | 36 (12.3%) | 13 (9.9%) | 49 (11.6%) |  |
| epsilon | 22 (7.5%) | 12 (9.2%) | 34 (8.0%) |  |
| omicron | 35 (12.0%) | 19 (14.5%) | 54 (12.8%) |  |
| **Primary vaccination completed** | 15 (5.1%) | 16 (12.2%) | 31 (7.3%) | .01 |
| **Symptoms** |  |  |  |  |
| Fatigue | 187 (64.0%) | 58 (44.3%) | 245 (57.9%) | <.001 |
| Shortness of breath | 146 (50.0%) | 42 (32.1%) | 188 (44.4%) | <.001 |
| Dizziness | 64 (21.9%) | 22 (16.8%) | 86 (20.3%) | .23 |
| Brain fog | 127 (43.5%) | 38 (29.0%) | 165 (39.0%) | .005 |
| Impaired memory | 99 (33.9%) | 28 (21.4%) | 127 (30.0%) | .009 |
| Headache | 96 (32.9%) | 36 (27.5%) | 132 (31.2%) | .27 |

(a) Long covid impact missing for n=6 persistent and 1 remitted participant

We applied Cox regression to examine the association with likelihood of remission with age, gender, self-reported severity of initial PCC symptoms, and predominant circulating viral variant at time of infection (Figure 1). All of these features were significantly associated with odds of symptom resolution at subsequent visit – those who were younger, male, who reported less than moderate severity of PCC symptoms, and who were fully vaccinated prior to initial COVID infection were more likely to experience remission.

**Figure 1.**
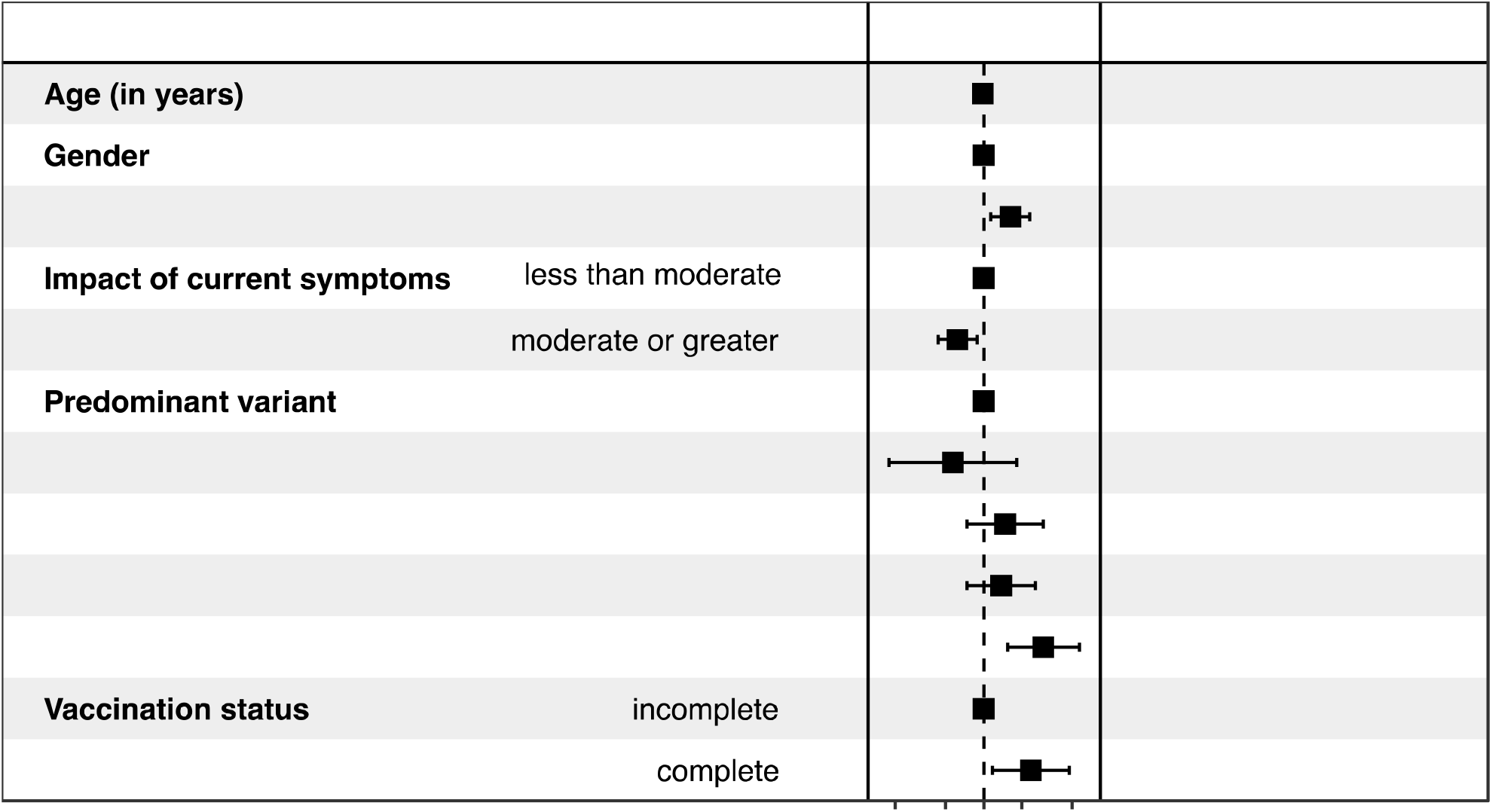
Forest plot of Cox proportional hazards regression model for remission hazard.

Figure 2 illustrates unadjusted differences in remission, by gender. Median time to remission among women respondents was 440 days (95% CI 349-556), compared to 316 (95% CI 210-411) among men; Kaplan-Meier log rank Chisq (1df) = 8.8, p= 0.003.

**Figure 2.**
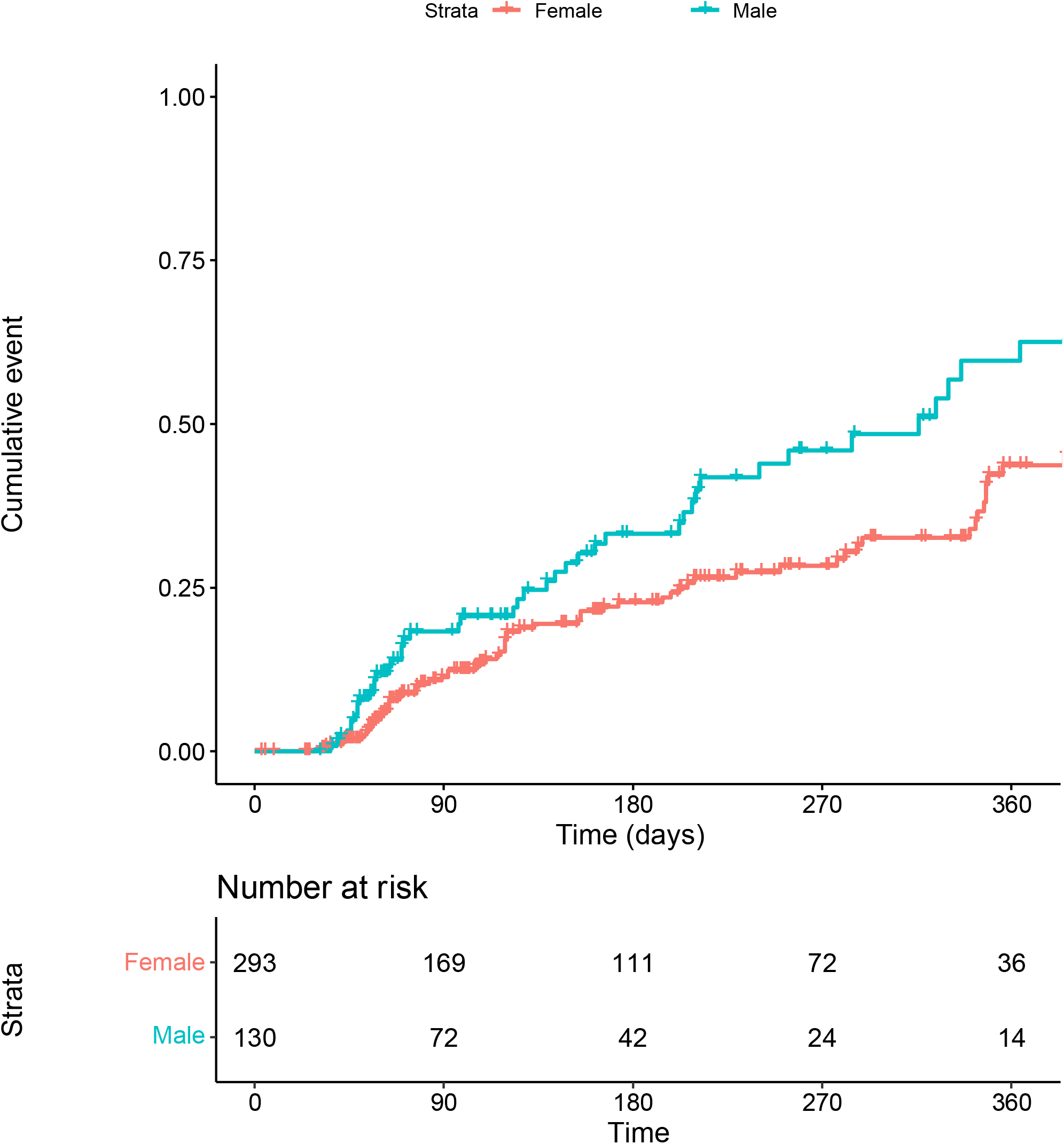
Kaplan-Meier survival curve examining time to remit among survey respondents identifying as male versus female.

We next examined associations between selected PCC symptoms present at initial survey and likelihood of remission, adjusted for age and gender (Figure 3). Both shortness of breath and brain fog, but not memory impairment, fatigue, or dizziness, were statistically significantly associated with diminished likelihood of remission. Figure 4 illustrates Kaplan-Meier curve for presence or absence of brain fog during initial survey; median time to remit was 348 days (95% CI 289-411) with the symptom absent, and 464 days (95% CI 385-NA) with the symptom present; log rank Chisq (1df) = 9.6 (1df), p= 0.002.

**Figure 3.**
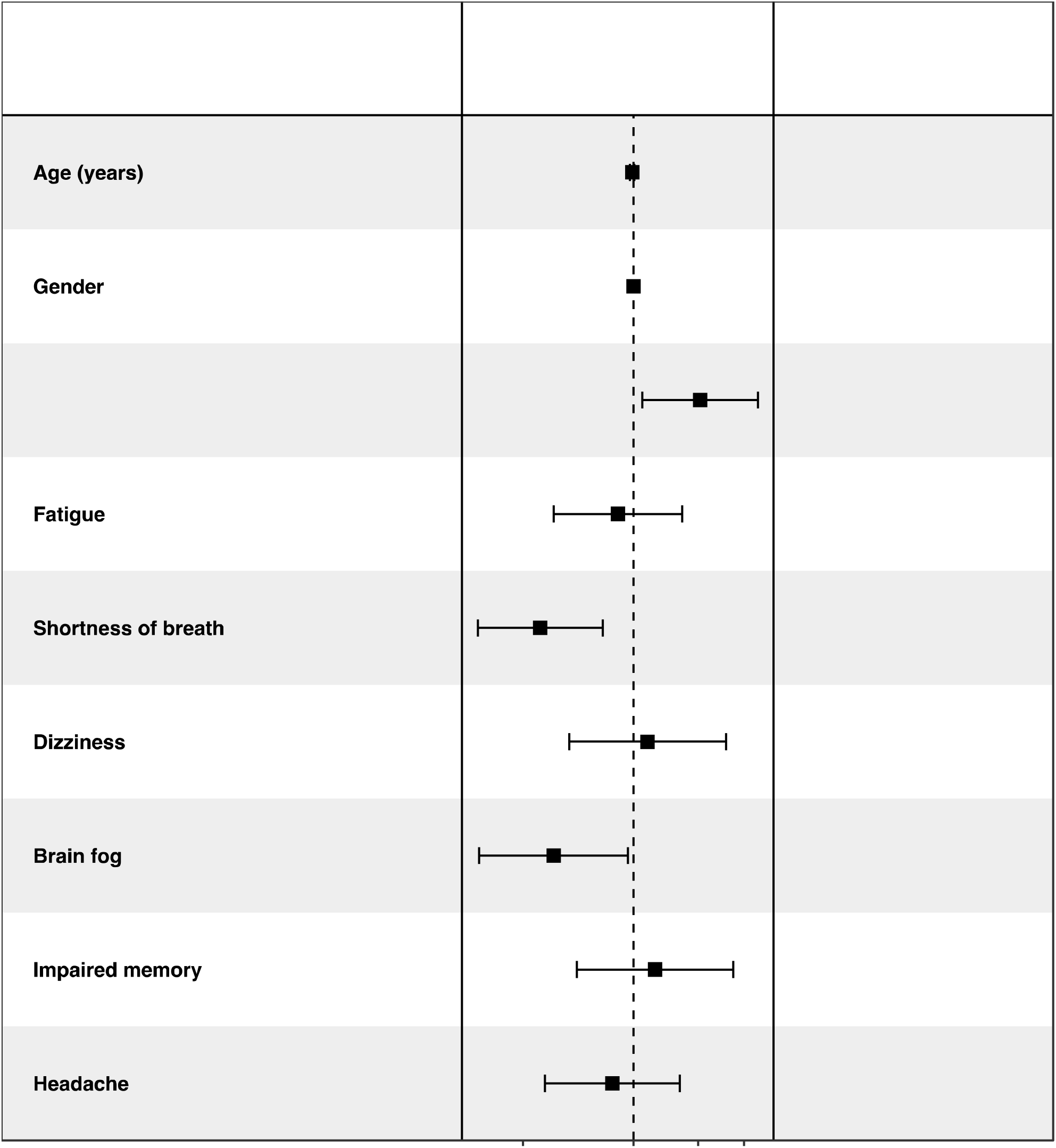
Forest plot of Cox proportional hazards regression model for remission hazard, examining selected symptoms present or absent at initial survey

**Figure 4.**
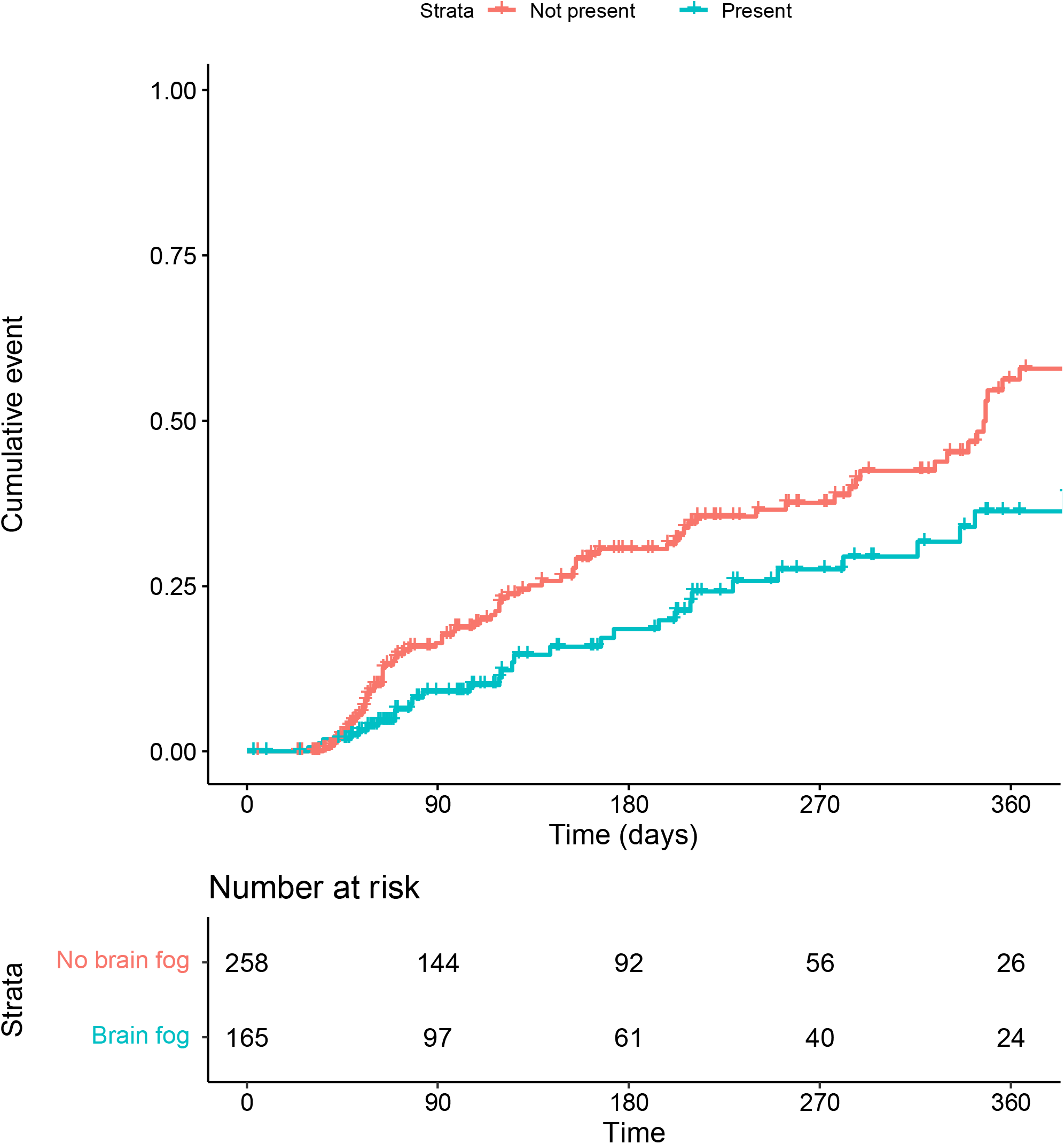
Kaplan-Meier survival curve examining association between presence or absence of ‘brain fog’ at initial survey and likelihood of subsequent remission

## DISCUSSION

In a cohort of individuals reporting symptoms of PCC surveyed at least twice, around 1/3 experienced remission during follow-up. Likelihood of remission was greatest among men, younger individuals, those who reported less severe PCC symptoms initially, and those infected when the Omicron strain predominated. Among individual symptoms of PCC, both brain fog and shortness of breath were associated with diminished likelihood of remission.

Prior work has suggested that younger age and male gender are associated with greater risk of developing PCC^185^. A survey-based analysis also suggested that infection during Omicron predominance was associated with lower risk^5^. The present study extends that literature by suggesting that risk factors for PCC itself may also represent risk factors for greater chronicity.

Two prior studies found lesser likelihood of remission among individuals with PCC. The first used a daily self-report symptom diary to estimate that about 15% of those who initially reported PCC had fully remitted at 1 year after acute infection^12^. A smaller 12-month follow-up survey study similarly estimated prevalence of remission of 9%^13^. Conversely, we find rates of remission nearly twice as great. As all 3 studies adopt different designs and sampling frames, they are not directly comparable; all are vulnerable to biases introduced by incomplete follow-up. Still, in aggregate they suggest that a subset of those who initially experience chronic symptoms will go on to remit, raising the central question of how to identify such individuals.

Prior work has suggested that younger age and male gender are associated with greater risk of developing PCC^185^. A survey-based analysis also suggested that infection during Omicron predominance was associated with lower risk^5^. The present study extends that literature by suggesting that risk factors for PCC itself may also represent risk factors for greater chronicity. In particular, the observation that vaccination prior to infection not only associated with diminished risk of developing PCC, but with likelihood of remission, provides yet another reason to emphasize prevention of acute infection wherever possible.

The pathophysiology of PCC remains poorly understood, with accumulating evidence of long-lasting immune and inflammatory dysregulation in a subset of individuals (for a review, see Mehandru^19^). This paucity of mechanistic understanding led to a lag in the initiation of treatment studies^11^, despite the prevalence of the syndrome^5^ and its associated disability. With the acceleration of such studies, the ability to identify individuals least likely to improve spontaneously may be valuable in maximizing the risk/benefit ratio of novel interventions.

Our identification of particular symptoms that predict poorer outcomes also merits further investigation. Notably, these symptoms do not reflect a single organ system, suggesting that they may instead relate to a common underlying process with multiple targets.

### Limitations

Our study has multiple limitations. The low number of repeat survey respondents may introduce biases; those who returned were older, more likely to be male, more highly educated, and more likely to be White, such that the generalizability of our results requires confirmation in other designs. The retrospective nature of recall (e.g., for dates of infection and vaccination) also may diminish reliability of our results. As a broad survey rather than one specifically aimed at characterizing illness in detail, we also lack measures of clinical detail that could also represent important predictors of remission, such as medical comorbidity, severity of initial infection, and number of infections. Many of these limitations will ultimately be addressed with prospective investigations such the NIH Recover-COVID study, with systematic and objective measurement of sequelae^20^, although some degree of attrition is likely to be unavoidable even with these designs.

### Conclusion

In spite of these caveats, this analysis indicates that a substantial proportion of individuals experience remission of PCC symptoms. Beyond the hopeful observation that recovery is possible, these results underscore the need to identify treatments that can increase remission and maximize function. While the potential impacts of PCC are only beginning to be described^98^, it is already apparent that large numbers of people will experience long-term consequences of their illness. By identifying those individuals least likely to experience spontaneous remission, these findings may help to target interventions to individuals most likely to benefit from them.

## Data Availability

Data are not yet available for distribution

## Acknowledgements

The survey was supported in part by the National Science Foundation (Drs. Ognyanova and Lazer) and by the National Institute of Mental Health (Drs. Perlis and Lazer - RF1MH132335). The sponsors did not have any role in design and conduct of the study; collection, management, analysis, and interpretation of the data; preparation, review, or approval of the manuscript; and decision to submit the manuscript for publication. The authors had the final responsibility for the decision to submit for publication. **Access to Data:** Dr. Perlis had full access to all the data in the study and takes responsibility for the integrity of the data and the accuracy of the data analysis.

## Author Contributions

Roy H. Perlis, MD MSc -- analyzed data, drafted and revised manuscript

Mauricio Santillana, PhD – revised manuscript

Katherine Ognyanova, PhD – generated data and contributed to analysis

David Lazer, PhD – revised manuscript

