## Supplemental Materials for "Correlates of symptomatic remission among individuals with post-COVID-19 condition"

Supplemental Table 1. Comparison of individuals who did or did not return following initial long COVID survey visit

|  | No return (N=3108) | Return  (N=423) | Total  (N=3531) | P value |
| --- | --- | --- | --- | --- |
| **Gender (n %)** |  |  |  | .005 |
| Female | 2350 (75.6%) | 293 (69.3%) | 2643 (74.9%) |  |
| Male | 758 (24.4%) | 130 (30.7%) | 888 (25.1%) |  |
| **Respondent age (mean SD)** | 42.5 (14.8) | 53.7 (13.6) | 43.8 (15.1) | <.001 |
| **Race and ethnicity** |  |  |  | <.001 |
| Black | 269 (8.7%) | 29 (6.9%) | 298 (8.4%) |  |
| Asian American | 80 (2.6%) | 9 (2.1%) | 89 (2.5%) |  |
| Hispanic | 327 (10.5%) | 13 (3.1%) | 340 (9.6%) |  |
| Native American | 32 (1.0%) | 2 (0.5%) | 34 (1.0%) |  |
| Other | 64 (2.1%) | 10 (2.4%) | 74 (2.1%) |  |
| Pacific Islander | 41 (1.3%) | 3 (0.7%) | 44 (1.2%) |  |
| White | 2295 (73.8%) | 357 (84.4%) | 2652 (75.1%) |  |
| **US region** |  |  |  | .22 |
| Midwest | 795 (25.6%) | 123 (29.1%) | 918 (26.0%) |  |
| Northeast | 426 (13.7%) | 62 (14.7%) | 488 (13.8%) |  |
| South | 1256 (40.4%) | 167 (39.5%) | 1423 (40.3%) |  |
| West | 631 (20.3%) | 71 (16.8%) | 702 (19.9%) |  |
| **Impact of long COVID symptoms**^a^ |  |  |  | .30 |
| Less than moderate | 1330 (43.2%) | 191 (45.9%) | 1521 (43.5%) |  |
| Moderate or greater | 1748 (56.8%) | 225 (54.1%) | 1973 (56.5%) |  |
| **Predominant COVID variant** |  |  |  | .12 |
| alpha | 173 (5.6%) | 24 (5.7%) | 197 (5.6%) |  |
| ancestral | 1794 (57.7%) | 262 (61.9%) | 2056 (58.2%) |  |
| delta | 518 (16.7%) | 49 (11.6%) | 567 (16.1%) |  |
| epsilon | 251 (8.1%) | 34 (8.0%) | 285 (8.1%) |  |
| omicron | 372 (12.0%) | 54 (12.8%) | 426 (12.1%) |  |
| **Primary vaccination completed** | 246 (7.9%) | 31 (7.3%) | 277 (7.8%) | .67 |
| **Symptoms** |  |  |  |  |
| Fatigue | 1616 (52.0%) | 245 (57.9%) | 1861 (52.7%) | .02 |
| Shortness of breath | 1221 (39.3%) | 188 (44.4%) | 1409 (39.9%) | .04 |
| Dizziness | 656 (21.1%) | 86 (20.3%) | 742 (21.0%) | .71 |
| Brain fog | 1845 (59.4%) | 258 (61.0%) | 2103 (59.6%) | .52 |
| Impaired memory | 869 (28.0%) | 127 (30.0%) | 996 (28.2%) | .38 |
| Headache | 1094 (35.2%) | 132 (31.2%) | 1226 (34.7%) | .11 |

a. Symptom severity missing for 30 nonreturning and 7 returning participants
